# The Gwangju Alzheimer’s & Related Dementias (GARD) Cohort: Over a Decade of Korea’s Largest Longitudinal Multimodal Study

**DOI:** 10.1101/2025.08.21.25333554

**Authors:** Kyu Yeong Choi, Sarang Kang, Seungho Cook, Donghe Li, Yu Yong Choi, Eun Hyun Seo, Xudong Han, Jung Eun Park, Suyeon Lee, Sunjae Lee, Ji Yeon Chung, Ari Chong, Seong-Min Choi, Jung-Min Ha, Min Kyung Song, Jung Sup Lee, IL Han Choo, Byeong C Kim, Hoowon Kim, Lindsay A. Farrer, Jungsoo Gim, Gyungah R. Jun, Kun Ho Lee

## Abstract

**INTRODUCTION:** Alzheimer’s disease (AD) is a major public health concern in Korea, with a high prevalence among older adults. A community-based longitudinal study is essential for tracking disease progression, identifying biomarkers, and developing targeted prevention and treatment strategies. The Gwangju Alzheimer’s and Related Dementias (GARD) cohort was established to address these needs through a multimodal approach.

**METHODS:** Participants aged ≥60 undergo comprehensive clinical evaluations, neuroimaging, and biospecimen collection for multi-omics analyses (genomics, transcriptomics, proteomics, microbiome) at baseline and systematic follow-up visits.

**RESULTS:** From over 17,000 screened individuals, 12,877 were enrolled. Baseline diagnoses include 5,995 cognitively unimpaired (CU), 4,025 mild cognitive impairment (MCI), and 1,026 AD dementia. The resource includes MRI scans (*n*=10,843) and extensive multi-omics data: genomic (*n*=10,775), proteome (*n*=116), and microbiome (*n*=595).

**DISCUSSION:** The integrated GARD dataset provides a powerful and scalable resource for identifying novel biomarkers, understanding disease heterogeneity, and advancing precision medicine for AD.

## 1. BACKGROUND

Alzheimer’s disease (AD) is a progressive neurodegenerative disorder and the most common cause of dementia worldwide. AD and related dementias represent a significant and escalating public health challenge in South Korea. Recent nationwide epidemiological studies have consistently documented a high prevalence and a continuous increase in the annual incidence of dementia among the old population, which the number of South Korean adults aged 65 and older is projected to surge from 8.53 million in 2021 to over 19 million by 2050 [1–3]. Beyond the clinical impact, the escalating number of cases imposes a substantial and growing economic burden on the nation’s healthcare system and society at large [4]. This pressing epidemiological and economic landscape underscores the urgent need for large-scale, deeply phenotyped longitudinal studies to identify risk factors, discover early biomarkers, and develop effective interventions.

Longitudinal studies in European-ancestry populations, such as the Alzheimer’s Disease Neuroimaging Initiative (ADNI) and the UK Biobank, have been instrumental in defining the trajectory of AD [5, 6]. Through the integration of multi-modal data, these cohorts have established the utility of structural MRI for detecting characteristic brain atrophy [7], demonstrated the correlation between amyloid-beta deposition and cognitive decline [8], and propelled the development of promising plasma biomarkers like p-tau217 [9]. Furthermore, large-scale genomic data from resources like the UK Biobank have significantly advanced our understanding of AD’s genetic architecture [6]. However, given the known ancestral differences in genetic risk and environmental factors, it is critical to determine whether these biomarkers and progression models are directly applicable to Koreans. This requires the establishment of dedicated, large-scale Korean cohorts to validate these findings and identify population-specific factors that influence AD pathogenesis.

The genetic architecture of Alzheimer’s disease (AD) is complex, with the *apolipoprotein E* (*APOE*) ε4 allele being the most significant known risk factor across diverse ancestries [10]. However, its impact is not uniform across all populations. In Koreans, for instance, not only is the ε4 allele less frequent compared to European-ancestry populations [11, 12], but other genetic variants within the *APOE* locus itself have been shown to modulate AD risk [13]. Although recent genome-wide association studies (GWAS) and whole-genome sequencing (WGS) in Koreans have identified novel risk loci beyond the *APOE* region, much of the heritability remains unexplained. Therefore, large-scale genetic studies within deeply phenotyped Korean cohorts are essential to uncover the remaining genetic architecture and build a comprehensive model of AD risk and progression.

The Gwangju Alzheimer’s and Related Dementias (GARD) cohort was specifically designed to confront the growing dementia crisis in South Korea by creating a deeply phenotyped, longitudinal research platform. Established in 2013, it integrated extensive neuropsychological, neuroimaging (MRI, amyloid-PET), biospecimen, and multi-omics data to track disease from its earliest stages. Here, we report overall study design and available resources from the GARD to (i) characterize Korean-specific genetic risk profiles, (ii) map multimodal biomarker trajectories, and (iii) identify novel biomarkers for cognitive decline.

## 2. METHODS

### 2.1 Study Approval and Community-Based Screening

This study was approved by the Institutional Review Boards of Chosun University Hospital (IRB No. CHOSUN-2013-12-018-070 and CHOSUN 2019-10-022-025) and Chonnam National University Hospital (IRB No. CNUH-2019-279). All research methods were conducted by the Declaration of Helsinki and relevant guidelines and regulations. Written informed consent was provided by all volunteers and family members or authorized caregivers in the case of cognitively impaired patients.

The GARD cohort implemented a structured community-based screening process to identify eligible individuals for participation. This process was conducted in collaboration with Gwangju’s Dementia Prevention and Management Center and affiliated local healthcare institutions. Trained research nurses and clinical psychologists administered standardized assessments to evaluate cognitive function, functional ability, and overall medical health status. Before undergoing formal screening, subjects were assessed against inclusion and exclusion criteria to determine their eligibility. Individuals who met the inclusion criteria proceeded to cognitive testing, while those with conditions such as severe other neurological disorders, psychiatric illnesses, or functional impairments that could interfere with cognitive evaluation were excluded.

Eligible subjects underwent cognitive and functional screening, which included the Korean Mini-Mental State Examination (K-MMSE) [14] as a global cognitive assessment tool. In addition, the Subjective Memory Complaints Questionnaire (SMCQ) [15] and Korean Dementia Screening Questionnaire (KDSQ) [16] were used to assess self-reported and caregiver-reported cognitive concerns. Functional status was evaluated using the Korean Instrumental Activities of Daily Living (K-IADL) [17], and depressive symptoms were screened through the Geriatric Depression Scale-K (GDS-K) [18]. These assessments provided an initial classification of cognitive status and functional ability. Alongside cognitive and functional testing, comprehensive medical history and demographic data were collected to assess potential risk factors for cognitive impairment. This included information on hypertension, diabetes, cardiovascular diseases, cerebrovascular conditions, psychiatric history, sensory impairments (hearing and vision loss), alcohol consumption, and smoking habits. Additionally, physical health parameters, such as blood pressure, body mass index (BMI), and waist-to-hip ratio, were recorded to evaluate overall health status.

### 2.2 Neuropsychological Assessment, Magnetic Resonance Imaging Scans, and Clinical Diagnosis

Following initial community-based screening, participants who met the eligibility criteria underwent in-depth neuropsychological evaluations and clinical diagnostic assessments to determine dementia status. These procedures were administered by trained neuropsychologists and dementia specialists, following standardized protocols to ensure diagnostic accuracy. Neuropsychological testing included the 2nd edition[19] of the Seoul Neuropsychological Screening Battery (SNSB)[20], evaluating five key cognitive domains: memory, executive function, attention, language, and visuospatial ability.

To comprehensively evaluate structural and functional brain changes associated with cognitive decline, the GARD cohort incorporated multimodal neuroimaging and electrophysiological assessments. High-resolution 3T magnetic resonance imaging (MRI) scans were acquired to assess brain morphology, structural integrity, and neurodegenerative changes. The MRI protocol included T1-weighted magnetization-prepared rapid gradient echo (T1 MPRAGE) for anatomical assessment, T2-weighted fluid-attenuated inversion recovery (T2 FLAIR) for detecting white matter hyperintensities, susceptibility-weighted imaging (SWI) for microbleed and iron deposition analysis, and diffusion tensor imaging (DTI) for white matter tract integrity evaluation. Functional neuroimaging was conducted using functional MRI (fMRI) to investigate resting-state and task-based functional connectivity patterns. This facilitated the identification of network dysfunction, particularly within the default mode network (DMN), which is known to be disrupted in AD progression.

Clinical diagnosis was based on cognitive test performance, functional ability, and medical history, following established guidelines such as the Clinical Dementia Rating (CDR), the Diagnostic and Statistical Manual of mental disorders, Fifth Edition (DSM-5) criteria for major neurocognitive disorder, and the National Institute on Aging and the Alzheimer’s Association (NIA-AA) research framework [21]. Participants were categorized into cognitively unimpaired (CU), mild cognitive impairment (MCI), or dementia due to AD based on this multi-source evaluation. To ensure diagnostic consistency, individuals with neurological, psychiatric, or medical conditions that could confound cognitive assessment—such as history of stroke, severe brain injury, epilepsy, or major depression—were excluded from CU classification. In addition, baseline magnetic resonance imaging (MRI) was used to screen for structural brain abnormalities incompatible with a CU diagnosis.

### 2.3 Amyloid PET Imaging and Electrophysiological Assessments

Amyloid positron emission tomography (PET) imaging using 18F-Florbetaben was performed to quantify β-amyloid deposition, with standardized uptake value ratio (SUVr) calculations used to determine brain amyloid plaque load (BAPL). Participants were categorized into negative, moderate, or severe amyloid burden groups, aiding in disease staging. Further, FDG-PET imaging was conducted to assess glucose metabolism as a marker of synaptic dysfunction, a characteristic of early AD. In addition to amyloid and FDG-PET imaging, CIT-PET was performed in a subset of participants to assess dopamine transporter availability, providing insight into dopaminergic system function and its relationship with cognitive decline.

Electrophysiological assessments including electroencephalography (EEG) and event-related potentials (ERP) were used to evaluate cortical activity and cognitive processing deficits. EEG recordings captured neural oscillations and abnormalities in resting-state brain activity, while ERP measurements provided insights into cognitive responses to stimuli, reflecting impairments in information processing and memory function. These assessments were served as functional biomarkers for detecting early neurophysiological changes in cognitive impairment.

### 2.4 Biospecimen Collection and Processing

Blood samples were obtained from participants to assess a range of hematological, metabolic, and biochemical markers associated with cognitive function and neurodegenerative disease. Fasting blood samples were collected into plastic collection tubes containing ethylenediamine tetra-acetic acid (EDTA) for the plasma and serum separator tube (SST) tube for serum. Samples were centrifuged twice at 2,000 x g and 3,000 x g at 4, and only the supernatant was extracted. The concentrated band of white blood cells, known as a buffy coat, was collected and used for DNA extraction. All the samples were aliquoted into 350ul and 165ul and stored in a -80 degrees deep freezer and a -196 degrees liquid nitrogen tank.

### 2.5 Laboratory Assessments and Fluid Biomarker Analysis

Routine blood tests included ABO blood type, white blood cell (WBC) count, red blood cell (RBC) count, platelet (PLT) count, hemoglobin (HGB), and hematocrit (Hct). Additionally, metabolic and biochemical markers were assessed, including fasting glucose, total cholesterol, low-density lipoprotein cholesterol (LDL), high-density lipoprotein cholesterol (HDL), triglycerides, glycated hemoglobin (HbA1c), creatinine, and thrombocyte levels. To assess systemic inflammation and immune responses, neutrophil segmentation (NeutrophilSeg), lymphocyte, monocyte, eosinophil, and basophil counts were obtained. Liver function tests including aspartate aminotransferase (AST/GOT) and alanine aminotransferase (ALT/GPT) were monitored for their potential associations with metabolic dysfunction and neurodegenerative processes [22].

AD-related plasma biomarkers were measured including phosphorylated Tau at 217 (pTau217), glial fibrillary acidic protein (GFAP), neurofilament light chain (NfL), and Aβ42/40, using the Simoa assay to capture subtle changes in neurodegeneration, glial activation, and amyloid pathology. Additionally, cerebrospinal fluid (CSF) samples were collected by a neurologist at Chonnam National University Hospital. The collection and storage procedures were described in previous studies [23]. The concentrations of Aβ42, Aβ40, total Tau, and pTau181 in CSF were measured using two different immunoassay platforms, INNOTEST ELISA and INNOBIA AlzBio3 xMAP (Fujirebio, Ghent, Belgium) platform [23, 24].

### 2.6 Generation of Genetic and Multi-Omics Data

The GARD cohort generated genetic, genomic, transcriptomic, proteomic, and metagenomic data for participants to identify genetic risk variants, transcriptomic signatures, and microbial compositions associated with AD susceptibility and progression in the Korean population. GWAS data were generated using the Affymetrix KNIH Biobank Array (v1.0 & v1.1) by the genome-wide single nucleotide polymorphism (SNP) array platform [25]. Whole-genome sequencing (WGS) data were obtained from the Illumina NovaSeq 6000 platform [26].

RNA sequencing (RNA-seq) data from blood were generated to investigate gene expression changes for AD and AD-related phenotypes. RNA-seq libraries were prepared using the TruSeq Stranded Total RNA Sample Prep Kit with Ribo-Zero H/M/R and sequenced on the NovaSeq 6000 platform, yielding high-resolution transcriptomic data. Proteomic profiling in plasma was conducted using a high-throughput aptamer-based technology (SomaScan 11K platform), enabling quantification of over 10,000 circulating proteins in plasma [27]. This approach provided a broad systems-level snapshot of biological pathways and novel biomarker detection for AD. A total of 2,214 plasma samples were measured to quantify pTau217, GFAP, and NfL, using ultra-sensitive immunoassays. Additionally, a total of 650 CSF samples were used to assess Aβ40, Aβ42, pTau181, and tTau, providing complementary molecular signatures of amyloid and tau pathology.

### 2.7 Generation of shotgun metagenomics data

Shotgun metagenomics of fecal, saliva, and dental samples were generated from the recruited participants. For saliva and dental samples, genomic DNA was Exgene kit (GeneAll) and for fecal samples, genomic DNA was extracted by DNeasy PowerSoil kit (Qiagen). Genomic DNA libraries of the all the given samples were prepared by the EZ-Tera XT DNA Library Prep Kit (Enzynomics, Cat. EZ036) according to the manufacturer’s protocol. Libraries were generated through a tagmentation process using transposome enzymes, which simultaneously fragment the DNA and attach adapter sequences. Briefly, 1–20 ng of input DNA was subjected to tagmentation using Tagment DNA Buffer and Amplicon Tagment Mix. Following the neutralization step, the tagmented DNA was amplified using indexed primers. The quality of the libraries was assessed using the TapeStation 4200 system and D5000 ScreenTape (Agilent Technologies, CA, USA). Libraries were quantified using the KAPA Library Quantification Kit (KK4824; Kapa Biosystems, MA, USA) according to the manufacturer’s protocol. Sequencing was performed as paired-end reads (2 × 150 bp) on the Illumina NovaSeq 6000 and X Plus platform (Illumina, CA, USA) following cluster amplification of denatured templates.

## 3. RESULTS

### 3.1 Cohort Recruitment and Baseline Characteristics

The GARD cohort was established in 2013 as a large-scale, community-based prospective study to investigate early risk and protective factors across the AD spectrum. **Figure 1** illustrated the overarching study framework (**Figure 1A**) and timeline (**Figure 1B**), outlining how participants have been tracked longitudinally across stages of cognitive change from cognitively unimpaired aging to asymptomatic AD, MCI, and eventual dementia diagnosis (**Figure 1C**). The GARD cohort primarily focused on participants who were in CU or had MCI but exhibited underlying pathological changes assessed by amyloid PET imaging, representing an asymptomatic or preclinical stage of disease (**Figure 1A**). Timeline of data collection, including the introduction of neuropsychological testing, neuroimaging (MRI, fMRI, PET), genetic analyses (GWAS chip, WGS), biospecimen collection (CSF, blood, microbiome), and electrophysiological recordings (EEG, ERP), implemented sequentially from 2013 to 2025 (**Figures 1B and 1C**).

**Figure 1.**
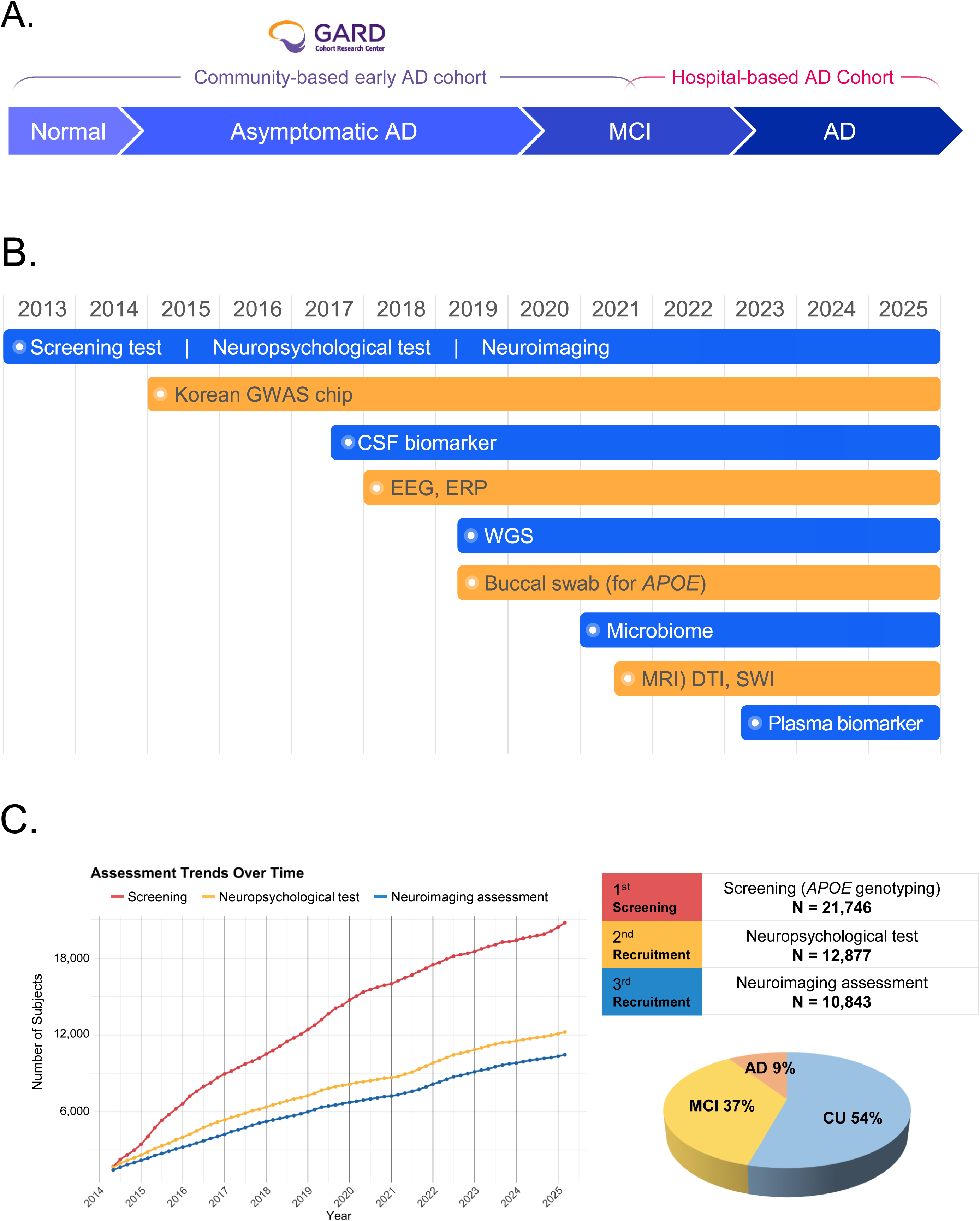
The GARD cohort study design and participant assessment workflow. **(A)** Overall study design. **(B)** Timeline of key assessments (neuropsychological, neuroimaging, genetic, and biomarker). **(C)** The three-stage screening process: (1) Initial Screening (MMSE, *APOE* genotyping); (2) Neuropsychological Testing; and (3) Neuroimaging (MRI).

Eligibility for advancement to the first stage of the GARD cohort was determined using a structured set of inclusion and exclusion criteria (**Table 1**). The inclusion criteria focused on recruiting community-dwelling older adults without a prior diagnosis of dementia or severe medical conditions. Exclusion criteria eliminated individuals with other neurological or psychiatric illnesses, uncontrolled chronic diseases, or sensory/motor impairments that could interfere cognitive testing. Based on these criteria, a total of 21,746 individuals were evaluated during the initial phase in 2013 and conducted brief cognitive tests including the K-MMSE and genetic risk profiling through *APOE* genotyping (**Figure 1C**). Individuals who were qualified for the study proceeded to comprehensively assessed multiple cognitive domains. Participants who completed this phase were officially enrolled in the study cohort.

**Table 1.** Overview of the GARD Cohort: Study Design, Inclusion/Exclusion Criteria, and FollowlUp Strategy.

| Cohort Name | Gwangju Alzheimer's & Related Dementias (GARD) Cohort |
| --- | --- |
| Study Type | Community-based longitudinal study |
| Study Period | 2013 – Present |
| Study Sites | Bitgoeul Senior Health Town, Chosun University Hospital, Chonnam National University Hospital |
| Inclusion Criteria | - Age ≥ 60 years<br>- No severe sensory impairments (hearing, vision, or language deficits affecting cognitive assessments)<br>- No major neurological or psychiatric disorders (stroke, epilepsy, schizophrenia, or substance use disorder)<br>- No significant physical disabilities or recent cancer diagnosis (past three years)<br>- No implanted metal devices (e.g., pacemakers, deep brain stimulators) that could interfere with MRI scans<br>- Willingness to provide consent and participate in follow-ups |
| Exclusion Criteria | - Severe sensory impairments affecting cognitive testing<br>- History of stroke, epilepsy, or major psychiatric disorders<br>- Severe physical disability or head trauma (with loss of consciousness)<br>- Cancer diagnosis within the last three years<br>- Presence of implanted metal devices (e.g., pacemakers, deep brain stimulators, metallic implants) that preclude MRI assessment<br>- Refusal to provide consent or participate in biospecimen collection |
| Follow-Up Criteria | - Amyloid PET-Positive: Follow-up every 1–1.5 years<br>- Amyloid PET-Negative: Follow-up every 1.5–2 years<br>- Borderline Amyloid (SUVR-A 1.3–1.39): Close monitoring every 1–1.5 years - *APOE* ε4 Carriers (no PET data): Annual follow-ups; PET scheduled as needed<br>- MRI Follow-Ups: Positive findings → Follow-ups synchronized with neuropsychological assessments; Negative findings → Follow-up only if cognitive decline is detected<br>- PET Follow-Ups: Amyloid-positive → Repeat PET every 3 years (unless BAPL indicates no further imaging is required); Amyloid-negative → Repeat PET every 3–5 years - Borderline Amyloid Cases: PET every 2 years to track amyloid progression and cognitive changes |

Abbreviations: **APOE**, apolipoprotein E **BAPL**, brain amyloid plaque load **MRI**, magnetic resonance imaging **PET**, positron emission tomography **SUVR**, standardized uptake value ratio

### 3.2 Neuroimaging Biomarker Assessment

Neuroimaging data were collected from all participants in the GARD cohort to facilitate accurate diagnosis and monitor the progression of brain atrophy (**Figure 1 and Table 2**). With over 13,000 structural MRI scans, the cohort enabled morphometric and volumetric analyses at a scale comparable to leading global imaging consortia. These were complemented by fMRI (n=8,579), DTI (n=3,717), and SWI (n=4,390), providing a robust framework for investigating network-level dynamics and microvascular changes. Notably, the acquisition of amyloid PET (n=5,800) and FDG-PET (n=446) scans in such a large sample is unprecedented among Asian cohorts, enabling molecular-level tracking of AD pathology with exceptional population coverage.

**Table 2.** Multimodal data in recruited GARD participants.

| Data Type and Category | Specific Measures / detail description | Total N |
| --- | --- | --- |
| **Neuropsychological test:** |  |  |
| SNSB | Memory, Language, Executive Function, Attention, Visuospatial | 12,877 |
| **Diagnosis:** |  |  |
| Prevalent Cases | Participants diagnosed with MCI or AD at baseline | CU: 5,995<br>MCI : 4,025<br>AD : 1,026<br>Others : 1,831 |
| Follow-Up & Incident Cases | Participants who progressed from CN or MCI to MCI or AD during follow-up | CU → MCI : 450<br>MCI → AD : 106<br>CU → AD : 17 |
| **Neuroimaging:** |  |  |
| Structural MRI | T1, T2, FLAIR | 10,843 |
| Functional MRI | Resting-State, Task-Based | 7,260 |
| Diffusion Tensor Imaging (DTI) | White Matter Integrity | 3,335 |
| Susceptibility Weighted Imaging (SWI) | Microbleeds, Iron Deposition | 3,839 |
| Amyloid PET | 18F-Florbetaben | 5,800 |
| FDG-PET | Glucose Metabolism in AD | 419 |
| **Electrophysiology:** EEG, ERP | Cortical Activity, Cognitive Processing | 7,060 |
| **Blood & Plasma:** |  |  |
| Vascular markers | Total cholesterol, HDL, LDL, Glucose, HbA1C, Creatinine, Thrombocyte | 10,639 |
| Plasma biomarker | pTau217, GFAP, NfL | 2,214 |
| **Cerebrospinal fluid (CSF) biomarker** | Aβ40, Aβ42, pTau181, tTau | 650 |
| **Genetic and omics:** |  |  |
| GWAS | Affymetrix KNIH biobank array | 12,877 |
| WGS | Illumina NovaSeq 6000 | 4,100 |
| Proteome | SomaScan 11k assay | 116 |
| RNASeq | NovaSeq 6000 | 600 |
| **Microbiome** |  |  |
| Gut microbiota | Feces | 595 |
| Oral microbiota | Saliva | 564 |
| Dental biofilm microbiota | Dental plaque | 405 |

Abbreviations: **SNSB**, Seoul Neuropsychological Screening Battery; **A $\beta$** , amyloid beta; **Aβ40 / Aβ42 / Aβ42/40**, isoforms of amyloid beta peptide; **AD**, Alzheimer's disease; **CU**, cognitively unimpaired; **Others**, included diagnosis is pending due to ongoing clinical assessment or other types of dementia; **CSF**, cerebrospinal fluid; **DTI**, diffusion tensor imaging; **EEG**, electroencephalography; **ERP**, event-related potential; **FDG**, fluorodeoxyglucose; **FLAIR**, fluid-attenuated inversion recovery; **fMRI**, functional magnetic resonance imaging; **GFAP**, glial fibrillary acidic protein; **GWAS**, genome-wide association study; **HbA1C**, glycated hemoglobin; **HDL**, high-density lipoprotein; **hs-CRP**, high sensitivity C-reactive protein; **IL-6**: interleukin-6; **LDL**, low-density lipoprotein; **MCI**, mild cognitive impairment; **MRI**, magnetic resonance imaging; **NfL**, neurofilament light chain; **PET**, positron emission tomography; **pTau181 / pTau217**, phosphorylated tau; **RNA-seq**, RNA sequencing; **SNSB**, Seoul Neuropsychological Screening Battery; **SWI**, susceptibility weighted imaging; **tTau**, total tau; **WGS**, whole genome sequencing

### 3.3 Diagnostic Classification and Longitudinal Trajectories

A total of 12,877 individuals underwent comprehensive neuropsychological assessments at baseline and follow-up, evaluating five cognitive domains (memory, language, executive function, attention, and visuospatial ability). The baseline cohort (mean age 72.17 ± 6.5 years; 62.98% female) exhibited wide cognitive variability (mean K-MMSE: 25.97 ± 3.7). Clinical diagnoses were determined using a multi-step algorithm (**Figure 2**). Participants were initially stratified by their CDR, with CDR=0 classified as CU. CU status required normal domain scores and no functional impairment, confirmed by excluding individuals with significant neuroimaging abnormalities (e.g., high WMH or MTA). Participants with mild cognitive deficits were diagnosed with MCI, while those with global cognitive and functional decline were classified as AD dementia, per NIA-AA guidelines.

**Figure 2.**
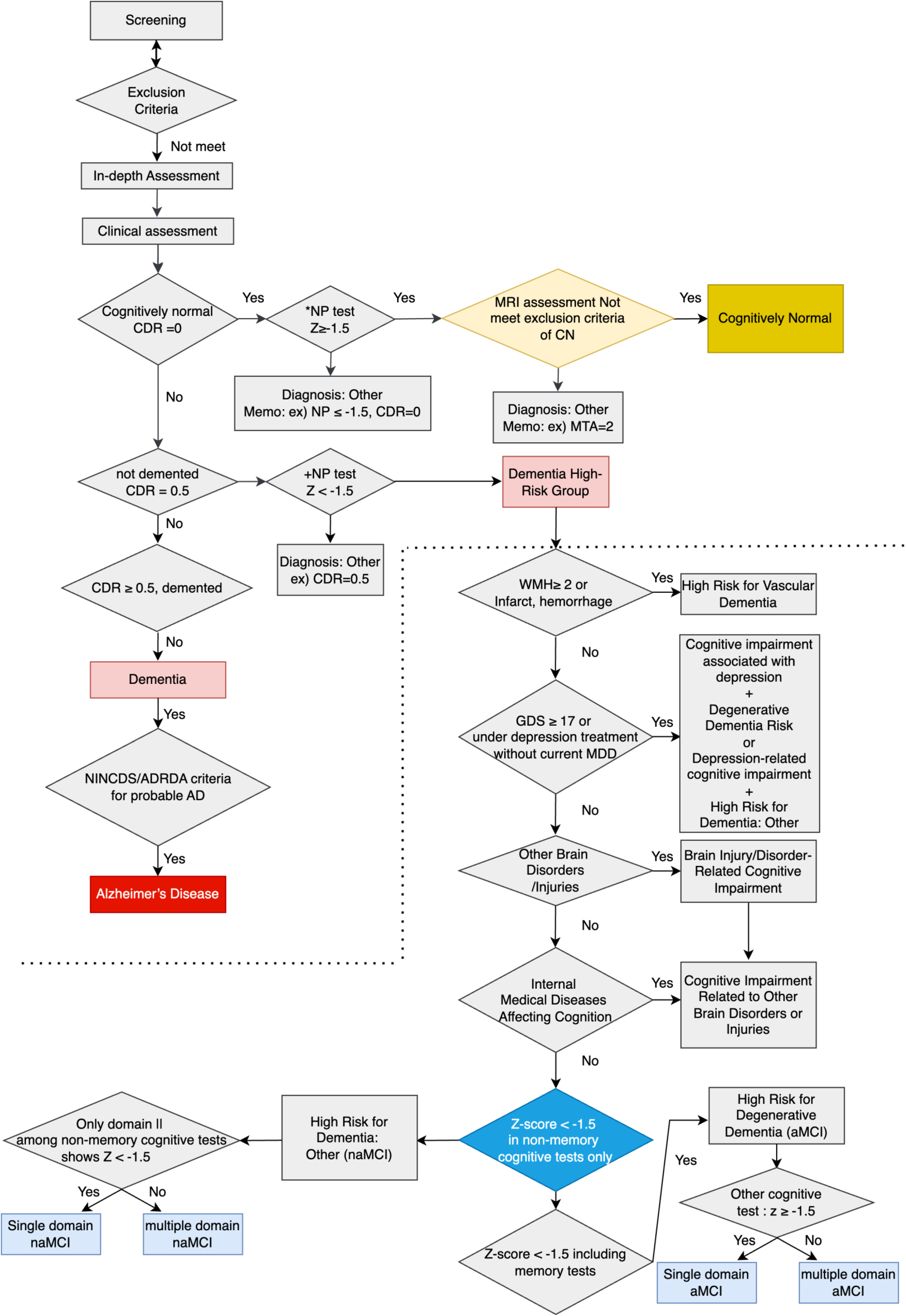
Flowchart of the diagnostic classification protocol for the GARD cohort. The figure details the sequential evaluation used to assign a diagnostic status. The process integrates initial eligibility and exclusion criteria with data from neuropsychological testing and neuroimaging. Final classification as Cognitively unimpaired (CU), Mild Cognitive Impairment (MCI), or Alzheimer’s Disease (AD) is determined by applying standardized thresholds, including Clinical Dementia Rating (CDR) scores.

At baseline, this process identified 4,025 participants with MCI and 1,026 with AD dementia (**Table 2**). A key feature of the GARD cohort is its biologically-informed follow-up strategy, which used clinical risk and amyloid PET results to create adaptive monitoring schedules. This approach was designed to identify individuals in the preclinical stage of AD, when pathology precedes symptoms. Over a decade, the cohort demonstrated significant diagnostic transitions, including 450 conversions from CU to MCI and 106 from MCI to AD, underscoring the heterogeneous trajectories of cognitive decline (**Table 2 and Figure 3**). The study’s robust longitudinal design is evidenced by 8,127 individuals completing at least one follow-up, with some completing up to nine assessments, enabling powerful tracking of long-term cognitive changes.

**Figure 3.**
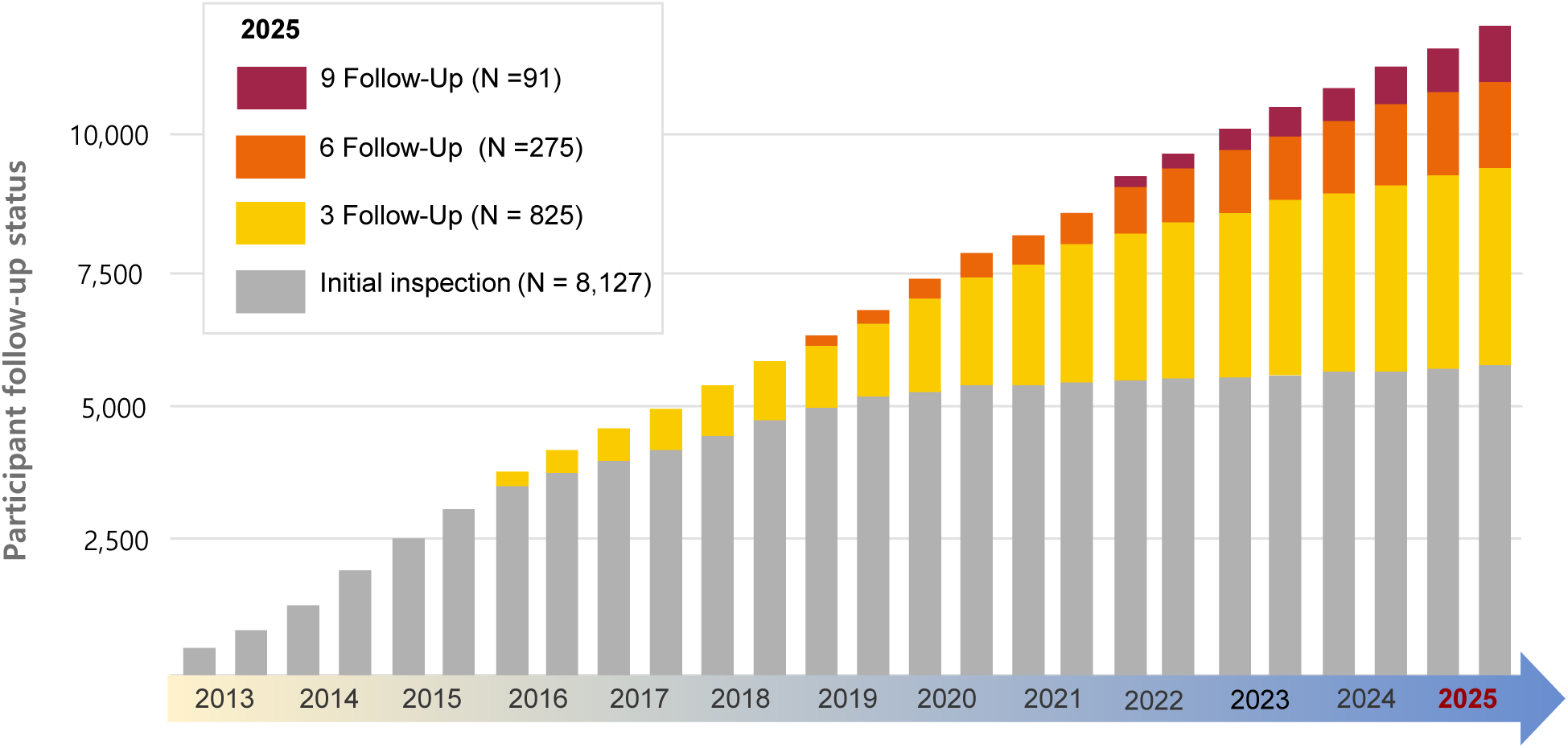
Longitudinal follow-up and diagnostic trajectories in the GARD cohort. This figure summarizes participant retention and illustrates the flow of individuals between cognitive states over time. It displays the distribution of participants by the number of completed follow-up assessments, highlighting the study’s long-term monitoring. The diagram also maps the trajectories of diagnostic stability and conversion between Cognitively unimpaired (CU), Mild Cognitive Impairment (MCI), and Alzheimer’s Disease (AD).

### 3.4 Comprehensive Multi-Modal Data Collection

The GARD cohort featured a comprehensive multimodal framework designed for integrative analysis of its extensive neuroimaging, physiological, and molecular data (**Figure 4 and Table 2**). Beyond the breadth of standard imaging (MRI, PET), a standout feature is the inclusion of 7,968 EEG/ERP scans, a scale rarely seen in population studies. This dataset provided a unique opportunity to investigate functional brain dynamics, such as network slowing, and their role in early cognitive decline.

**Figure 4.**
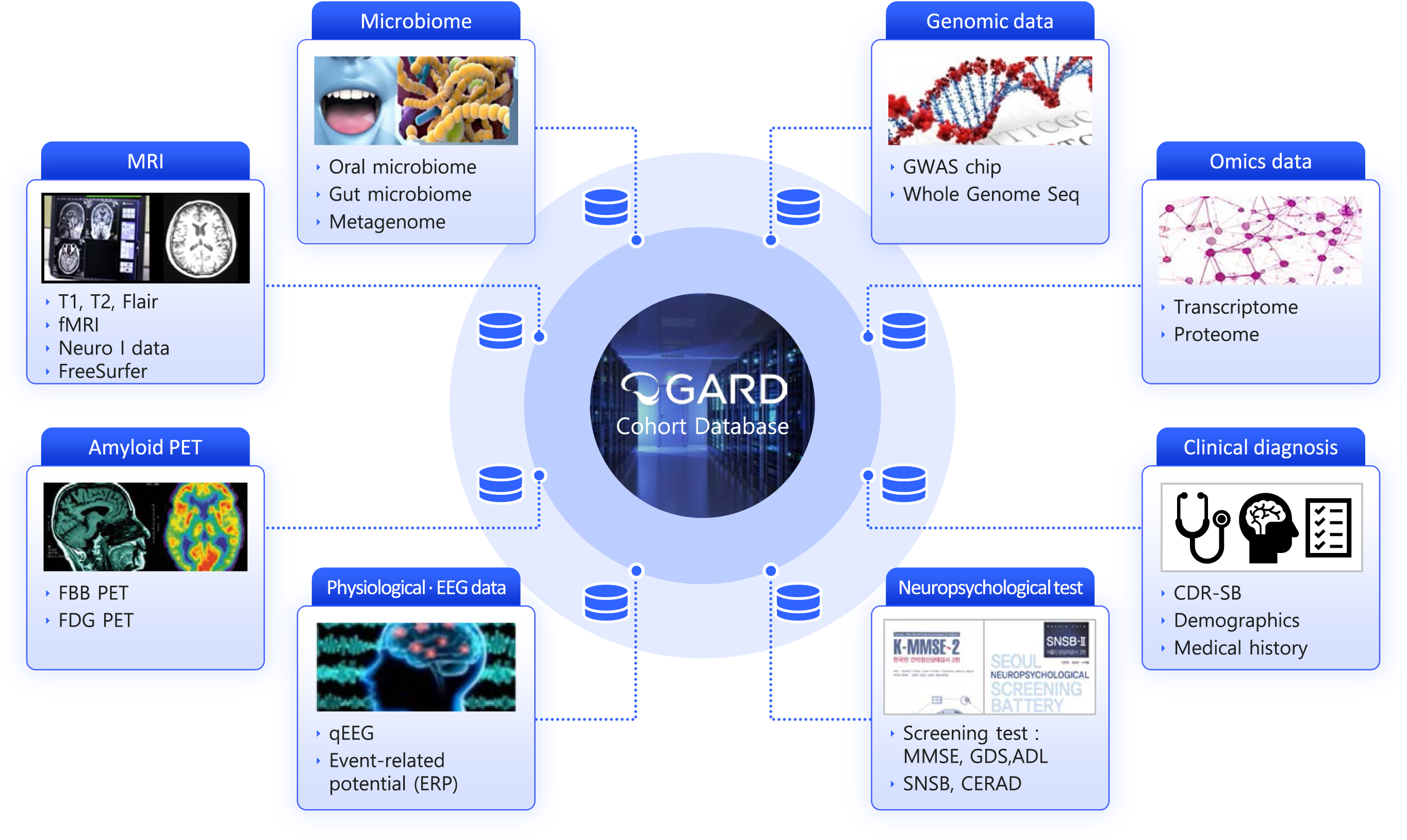
The GARD cohort multi-modal data platform. This diagram illustrates the comprehensive data platform, which integrates clinical and neuropsychological assessments with multi-omics, neuroimaging, and electrophysiological data. Key datasets include genomics, transcriptomics, proteomics, microbiome, neuroimaging (e.g., MRI), and electroencephalography (EEG). This multi-modal approach facilitates a systems-level investigation of disease progression and risk factors.

To complement the imaging data, the cohort included extensive biochemical and molecular markers. Routine blood sampling provided key biomarkers (e.g., lipids, glucose, creatinine) to assess cerebrovascular burden and its intersection with cognitive decline. Furthermore, to enable a systems-level understanding of AD pathophysiology, the study incorporated multi-omics datasets, including proteomics, transcriptomics, and epigenetics from blood samples.

### 3.4 Genetic Data Processing

The GWAS data were collected from three different batches: Blood Batch 1, Blood Batch 2, and Buccal. Each batch underwent standard downstream quality control procedures as described in a previous study [28]. Genotypes were excluded if they had a genotype call rate <95% or deviated from Hardy-Weinberg equilibrium (HWE) (p < 1×10). Individuals were excluded based on the following criteria: identity-by-state (IBS) >0.9 indicating potential duplicates; X-chromosome homozygosity between 0.2 and 0.8 or discordance with reported phenotypic sex; genotype call rate <95%; or SNP heterozygosity rate exceeding three standard deviations from the mean. To assess the potential batch effects across the three datasets, all samples were merged with the 1000 Genomes Project reference panel [29], and the clustering analysis was performed with principal components (PCs). Most subjects clustered with the Asian (ASN) population, and no batch effects were detected. Outliers and overlapping samples were removed based on the PCA clustering results. After the extensive quality control, a total of 12,877 subjects remained. Imputation was performed using the Trans-Omics for Precision Medicine (TOPMed) imputation server, applying the TOPMed version r3 reference panel [30]. Quality control followed the procedures described in [30], and imputed variants with an INFO score < 0.3 were excluded. After filtering 17,656,834 SNPs remained. Functional annotation of the variants was conducted using the ANNOVAR software. [31]

For the whole-genome sequencing (WGS) data, a total of 4,100 blood-derived DNA samples were submitted to DNA Link Inc. (Seoul, Republic of Korea) for sequencing and alignment. The sequencing and quality control procedures have been described in detail previously.[32] Following data processing, 49,074,372 SNPs, 2,488,596 insertions, and 3,390,751 deletions were retained for downstream analysis [32].

### 3.7 Microbiome Data Characteristics

Imbalances of human microbiome, called dysbiosis, have been associated with the progression of many chronic diseases, including neurodegenerative diseases like Parkinson’s diseases. To identify Alzheimer’s disease associated dysbiosis, we generated the shotgun metagenomics of fecal, saliva, and dental samples of the participants from the GARD cohort, totaling 1,564 shotgun metagenomic samples with replicates for some samples (595 fecal samples, 564 saliva samples, and 405 dental samples). First, we preprocessed raw sequencing reads of given metagenomic samples by illumina NovaSeq 6000 and X plus platform and removed low-quality reads and adapter sequences by AlienTrimmer (ver 2.1) [33]. Afterward, we extracted non-human reads from shotgun metagenomics data by kneaddata pipeline of bioBakery worfklow (ver 0.12.0) [34]. We used those high-quality reads for the compositional and functional profiling by bioBakery workflow, such as MetaPhlan4 [35] and HUMAnN3 [36].

## 4. DISCUSSION

The GARD cohort is a large-scale, prospective, population-based study of dementia in South Korea, uniquely designed to overcome the limitations of previous hospital-based (CREDOS [37]), clinic-recruited (KBASE [38]), or cross-sectional (KLOSCAD [39]) national studies. Its community-based recruitment and deep, longitudinal phenotyping allowed for unparalleled investigation into the natural history of cognitive decline. On a global scale, GARD addressed key gaps left by major cohorts. Unlike a general health platform in United Kingdom (UK Biobank; [6]) or FHS [40], the GARD provided harmonized, dementia-specific clinical and biomarker data. Unlike a memory clinic-based cohort in United States, National Alzheimer’s Coordinating Center (NACC; [41]), its population-level design captured the crucial preclinical stages of AD. These cohorts are predominantly European-ancestry populations. The Asian Cohort for Alzheimer’s Disease (ACAD) was developed to increase representation of Asian ancestry in dementia research, with a focus on Chinese, Korean, and Vietnamese adults in North America [42]. It integrates community-based recruitment as well as memory clinic-based recruitment strategies with multilingual cognitive assessment tools, genotyping, and plasma biomarker assessment. While ACAD is well-conceived in its scope and goals, it is still in its early phases. Neuroimaging, biospecimen collection, and longitudinal clinical follow-up are not yet fully implemented.

The GARD cohort’s primary strength is its design as a large-scale, prospective, population-based study tracking the full spectrum of cognitive aging in a real-world East Asian setting. Its harmonized infrastructure integrates deep, multimodal data, from clinical assessments and neuroimaging to molecular biomarkers, under a single, standardized protocol. A defining feature is its risk-stratified follow-up model, which enhances sensitivity to early disease transitions by frequent assessment of individual’s risk using cognitive assessments, neuroimaging scans, and fluid biomarker assays. This integrated, adaptive framework provides a vital and unique resource for global dementia research.

Despite these strengths, several limitations must be noted. The community-based recruitment, while ideal for studying early disease, results in fewer participants with moderate-to-severe AD and suffers lower number of AD or dementia cases at baseline and follow-ups. While data collection is extensive, certain advanced modalities (e.g., tau PET) and omics profiling are not yet universally available across the cohort. Finally, the adaptive follow-up schedule, though a strength for efficiency, creates variable temporal resolution across participants, which requires careful consideration in longitudinal analyses. Future directions aim to address these limitations directly. Priorities include expanding recruitment to include more individuals with advanced dementia and from diverse geographic regions. Broader implementation of advanced biomarker and omics platforms across the entire cohort is also essential. These steps will enhance GARD’s value as a comprehensive resource for translational research across the full continuum of AD.

In conclusion, the GARD cohort provides an essential and unique resource for understanding Alzheimer’s disease in a deeply phenotyped, population-based Korean cohort. Its novel design—combining community-based recruitment, harmonized multimodal data, and adaptive longitudinal follow-up—offers an unprecedented view into the preclinical and early symptomatic stages of dementia. By filling a critical demographic and methodological gap in global research infrastructure, GARD is positioned to drive significant advances in biomarker discovery, risk modeling, and the development of more inclusive and effective strategies to combat dementia worldwide.

## Data Availability

All data produced in the present study are available upon reasonable request to the authors

## Acknowledgements

We gratefully acknowledge the support of the Asia Dementia Foundation, Kolab Inc., and the members of the GARD cohort research group, whose commitment and collaboration have been essential to the development and implementation of this research.

## Consent Statement

All study participants, or their legal guardian, provided informed written consent prior to study enrollment.

## REFERENCES

[1] Baek MS, Kim HK, Han K, Kwon HS, Na HK, Lyoo CH, et al. Annual Trends in the Incidence and Prevalence of Alzheimer’s Disease in South Korea: A Nationwide Cohort Study. Front Neurol. 2022;13:883549.

[2] Jang JW, Park JH, Kim S, Lee SH, Lee SH, Kim YJ. Prevalence and Incidence of Dementia in South Korea: A Nationwide Analysis of the National Health Insurance Service Senior Cohort. J Clin Neurol. 2021;17:249–56.

[3] Shin JH. Dementia Epidemiology Fact Sheet 2022. Ann Rehabil Med. 2022;46:53–9.

[4] Shon C, Yoon H. Health-economic burden of dementia in South Korea. BMC Geriatr. 2021;21:549.

[5] Mueller SG, Weiner MW, Thal LJ, Petersen RC, Jack C, Jagust W, et al. The Alzheimer’s disease neuroimaging initiative. Neuroimaging clinics of North America. 2005;15:869–77, xi-xii.

[6] Bycroft C, Freeman C, Petkova D, Band G, Elliott LT, Sharp K, et al. The UK Biobank resource with deep phenotyping and genomic data. Nature. 2018;562:203–9.

[7] Frisoni GB, Fox NC, Jack CR, Jr., Scheltens P, Thompson PM. The clinical use of structural MRI in Alzheimer disease. Nat Rev Neurol. 2010;6:67–77.

[8] Villemagne VL, Burnham S, Bourgeat P, Brown B, Ellis KA, Salvado O, et al. Amyloid β deposition, neurodegeneration, and cognitive decline in sporadic Alzheimer’s disease: a prospective cohort study. The Lancet Neurology. 2013;12:357–67.

[9] Clark C, Lewczuk P, Kornhuber J, Richiardi J, Marechal B, Karikari TK, et al. Plasma neurofilament light and phosphorylated tau 181 as biomarkers of Alzheimer’s disease pathology and clinical disease progression. Alzheimers Res Ther. 2021;13:65.

[10] Rajabli F, Benchek P, Tosto G, Kushch N, Sha J, Bazemore K, et al. Multi-ancestry genome-wide meta-analysis of 56,241 individuals identifies LRRC4C, LHX5-AS1 and nominates ancestry-specific loci PTPRK, GRB14, and KIAA0825 as novel risk loci for Alzheimer’s disease: the Alzheimer’s Disease Genetics Consortium. medRxiv : the preprint server for health sciences. 2023.

[11] Kang S, Gim J, Lee J, Gunasekaran TI, Choi KY, Lee JJ, et al. Potential Novel Genes for Late-Onset Alzheimer’s Disease in East-Asian Descent Identified by APOE-Stratified Genome-Wide Association Study. J Alzheimers Dis. 2021;82:1451–60.

[12] Kang M, Farrell JJ, Zhu C, Park H, Kang S, Seo EH, et al. Whole-genome sequencing study in Koreans identifies novel loci for Alzheimer’s disease. Alzheimers Dement. 2024;20:8246–62.

[13] Choi KY, Lee JJ, Gunasekaran TI, Kang S, Lee W, Jeong J, et al. APOE Promoter Polymorphism-219T/G is an Effect Modifier of the Influence of APOE ε4 on Alzheimer’s Disease Risk in a Multiracial Sample. Journal of clinical medicine. 2019;8.

[14] Kim TH, Jhoo JH, Park JH, Kim JL, Ryu SH, Moon SW, et al. Korean version of mini mental status examination for dementia screening and its’ short form. Psychiatry Investig. 2010;7:102–8.

[15] Youn JC, Kim KW, Lee DY, Jhoo JH, Lee SB, Park JH, et al. Development of the subjective memory complaints questionnaire. Dementia and geriatric cognitive disorders. 2009;27:310–7.

[16] Yang D-W, Chey J-Y, Kim S-Y, Kim B-S. The development and validation of Korean dementia screening questionnaire (KDSQ). Journal of the Korean Neurological Association. 2002:135–41.

[17] Won CW, Yang KY, Rho YG, Kim SY, Lee EJ, Yoon JL, et al. The development of Korean activities of daily living (K-ADL) and Korean instrumental activities of daily living (K-IADL) scale. Journal of the Korean Geriatrics Society. 2002;6:107–20.

[18] Bae JN, Cho MJ. Development of the Korean version of the Geriatric Depression Scale and its short form among elderly psychiatric patients. Journal of psychosomatic research. 2004;57:297–305.

[19] Ryu HJ, Yang DW. The Seoul Neuropsychological Screening Battery (SNSB) for Comprehensive Neuropsychological Assessment. Dement Neurocogn Disord. 2023;22:1–15.

[20] Kang Y, Na D, Hahn SJIHbr, co c. Seoul neuropsychological screening battery. Incheon: Human brain research. 2003.

[21] Jack CR, Jr., Bennett DA, Blennow K, Carrillo MC, Dunn B, Haeberlein SB, et al. NIA-AA Research Framework: Toward a biological definition of Alzheimer’s disease. Alzheimers Dement. 2018;14:535–62.

[22] Weinstein G, Zelber-Sagi S, Preis SR, Beiser AS, DeCarli C, Speliotes EK, et al. Association of Nonalcoholic Fatty Liver Disease With Lower Brain Volume in Healthy Middle-aged Adults in the Framingham Study. JAMA Neurol. 2018;75:97–104.

[23] Lim HJ, Park JE, Kim BC, Choi SM, Song MK, Cho SH, et al. Comparison of Two Analytical Platforms in Cerebrospinal Fluid Biomarkers for the Classification of Alzheimer’s Disease Spectrum with Amyloid PET Imaging. J Alzheimers Dis. 2020;75:949–58.

[24] Park JE, Choi KY, Kim BC, Choi SM, Song MK, Lee JJ, et al. Cerebrospinal Fluid Biomarkers for the Diagnosis of Prodromal Alzheimer’s Disease in Amnestic Mild Cognitive Impairment. Dement Geriatr Cogn Dis Extra. 2019;9:100–13.

[25] Moon S, Kim YJ, Han S, Hwang MY, Shin DM, Park MY, et al. The Korea Biobank Array: design and identification of coding variants associated with blood biochemical traits. Scientific reports. 2019;9:1382.

[26] Modi A, Vai S, Caramelli D, Lari M. The Illumina sequencing protocol and the NovaSeq 6000 system. Bacterial pangenomics: methods and protocols: Springer; 2021. p. 15–42.

[27] Gold L, Ayers D, Bertino J, Bock C, Bock A, Brody EN, et al. Aptamer-based multiplexed proteomic technology for biomarker discovery. PLoS One. 2010;5:e15004.

[28] Anderson CA, Pettersson FH, Clarke GM, Cardon LR, Morris AP, Zondervan KT. Data quality control in genetic case-control association studies. Nat Protoc. 2010;5:1564–73.

[29] Genomes Project C, Auton A, Brooks LD, Durbin RM, Garrison EP, Kang HM, et al. A global reference for human genetic variation. Nature. 2015;526:68–74.

[30] Das S, Forer L, Schonherr S, Sidore C, Locke AE, Kwong A, et al. Next-generation genotype imputation service and methods. Nat Genet. 2016;48:1284–7.

[31] Wang K, Li M, Hakonarson H. ANNOVAR: functional annotation of genetic variants from high-throughput sequencing data. Nucleic Acids Res. 2010;38:e164.

[32] Kang M, Farrell JJ, Zhu C, Park H, Kang S, Seo EH, et al. Whole genome sequencing study in Koreans identifies novel loci for Alzheimer’s disease. Alzheimer’s & Dementia. 2024;20:8246–62.

[33] Criscuolo A, Brisse S. AlienTrimmer: a tool to quickly and accurately trim off multiple short contaminant sequences from high-throughput sequencing reads. Genomics. 2013;102:500–6.

[34] McIver LJ, Abu-Ali G, Franzosa EA, Schwager R, Morgan XC, Waldron L, et al. bioBakery: a meta’omic analysis environment. Bioinformatics. 2018;34:1235–7.

[35] Blanco-Míguez A, Beghini F, Cumbo F, McIver LJ, Thompson KN, Zolfo M, et al. Extending and improving metagenomic taxonomic profiling with uncharacterized species using MetaPhlAn 4. Nature biotechnology. 2023;41:1633–44.

[36] Beghini F, McIver LJ, Blanco-Míguez A, Dubois L, Asnicar F, Maharjan S, et al. Integrating taxonomic, functional, and strain-level profiling of diverse microbial communities with bioBakery 3. eLife. 2021;10.

[37] Park HK, Na DL, Han SH, Kim JY, Cheong HK, Kim SY, et al. Clinical characteristics of a nationwide hospital-based registry of mild-to-moderate Alzheimer’s disease patients in Korea: a CREDOS (Clinical Research Center for Dementia of South Korea) study. J Korean Med Sci. 2011;26:1219–26.

[38] Byun MS, Yi D, Lee JH, Choe YM, Sohn BK, Lee JY, et al. Korean Brain Aging Study for the Early Diagnosis and Prediction of Alzheimer’s Disease: Methodology and Baseline Sample Characteristics. Psychiatry Investig. 2017;14:851–63.

[39] Kim KW, Park JH, Kim MH, Kim MD, Kim BJ, Kim SK, et al. A nationwide survey on the prevalence of dementia and mild cognitive impairment in South Korea. J Alzheimers Dis. 2011;23:281–91.

[40] Mahmood SS, Levy D, Vasan RS, Wang TJ. The Framingham Heart Study and the epidemiology of cardiovascular disease: a historical perspective. Lancet. 2014;383:999–1008.

[41] Beekly DL, Ramos EM, Lee WW, Deitrich WD, Jacka ME, Wu J, et al. The National Alzheimer’s Coordinating Center (NACC) database: the Uniform Data Set. Alzheimer Dis Assoc Disord. 2007;21:249–58.

[42] Ho PC, Yu WH, Tee BL, Lee WP, Li C, Gu Y, et al. Asian Cohort for Alzheimer’s Disease (ACAD) pilot study on genetic and non-genetic risk factors for Alzheimer’s disease among Asian Americans and Canadians. Alzheimers Dement. 2024;20:2058–71.

